# Effects of Theophylline on ADCY5 Activation - From Cellular Studies to Improved Therapeutic Options for ADCY5-Related Dyskinesia Patients

**DOI:** 10.1101/2022.05.24.22274941

**Authors:** Dirk Tänzler, Marc Kipping, Marcell Lederer, Wiebke F. Günther, Christian Arlt, Stefan Hüttelmaier, Andreas Merkenschlager, Andrea Sinz

## Abstract

We show the effects of the three purine derivatives, caffeine, theophylline, and istradefylline, on the cAMP production of adenylyl cyclase 5 (ADCY5)-overexpressing cell lines. A comparison of cAMP levels was performed for ADCY5 wild-type and R418W mutant cells. ADCY5-catalyzed cAMP production was reduced with all three purines, while the most pronounced effects on cAMP reduction were observed for ADCY5 R418W mutant cells. The gain-of-function ADCY5 R418W mutant is characterized by an increased catalytic activity resulting in elevated cAMP levels, resulting in kinetic disorders or dyskinesia in patients. Based on our findings in ADCY5 cells, a slow-release formulation of theophylline was administered to a preschool aged patient with ADCY5-related dyskinesia. A striking improvement of symptoms was observed, outperforming the effects of caffeine that had previously been administered to the same patient.

## Background

Adenylyl cyclases (ADCYs) are central enzymes in all organisms (1). The ADCY family comprises ten isoforms; nine of them are membrane-bound enzymes (2,3). ADCY5 is one of the least studied isoforms that is most commonly expressed in brain and heart tissue (3). As all ADCYs, ADCY5 converts adenosine triphosphate (ATP) to cyclic adenosine-3’,5’-monophosphate (cAMP) and pyrophosphate (Figure 1) (3). ADCY5 has been identified as the primary ADCY isoform that is responsible for up to 80% of the cAMP production in striatal medium spiny neurons (3,4). The complex system of initiating and controling movement is dependent on a well-balanced signaling between G-Protein-Coupled-Receptors (GPCRs) and ADCYs (4). Different neurotransmitters stimulate or inhibit hydrolysis of ATP to cAMP via ADCY-coupled GPCRs (5). As such, adenosine receptor 2A (A_2A_) agonists increase intercellular cAMP levels (5).

**Figure 1.**
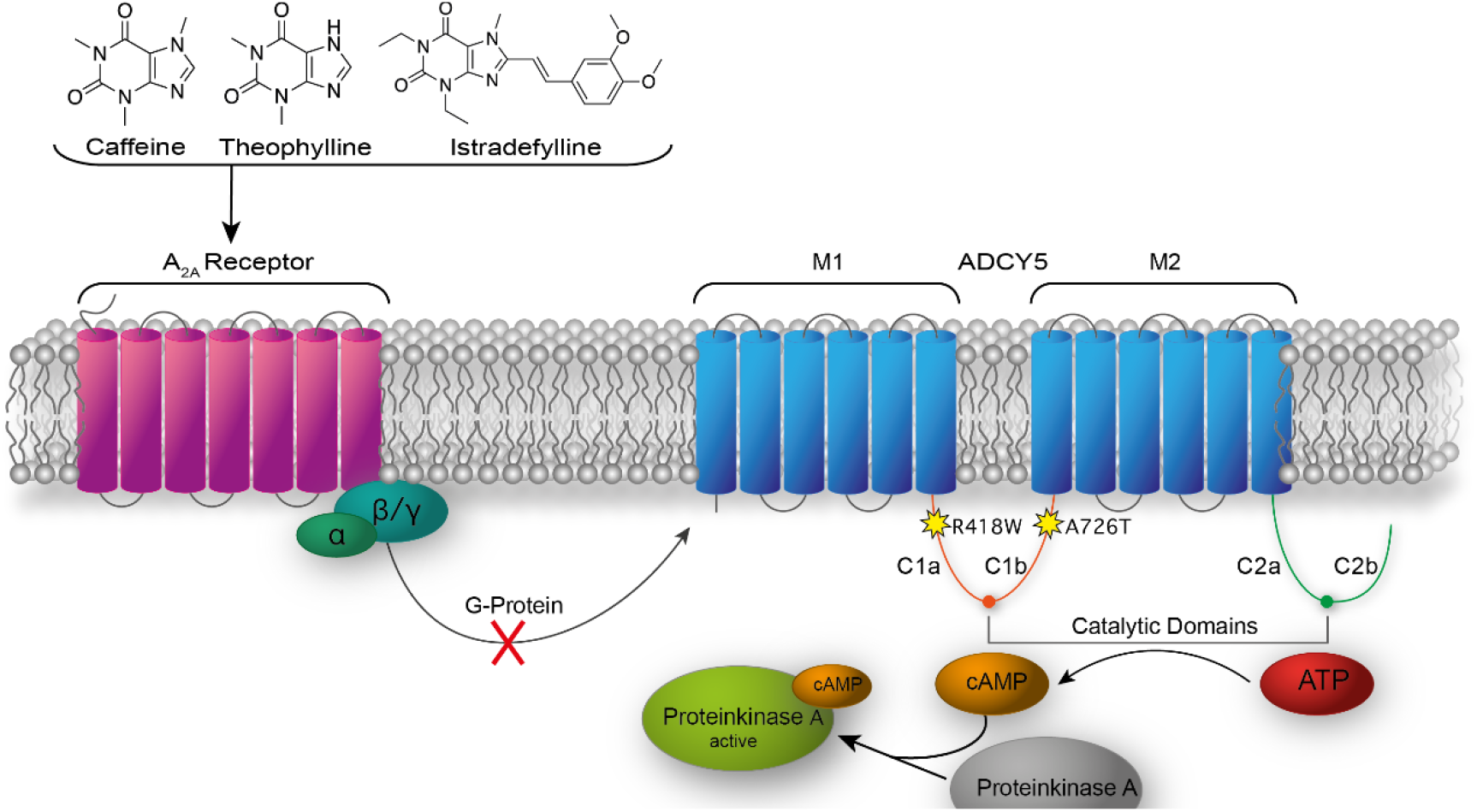
Mode of action for caffeine, theophylline, and istradefylline on the reduction of cAMP production by ADCY5 via inhibiting the A_2A_ receptor.

ADCY5 contains an intercellular N-terminal domain, two membrane domains (M1 and M2) consisting of six helices, two cytoplasmic homologous domains (C1 and C2), and a short intracellular loop between the two catalytic domains (Figure 1) (2,3,6). Crucial for ADCY5’s catalytic activity is the formation of an ATP-binding pocket, bringing together the C1 and C2 domains (2,3). Mutations in ADCY5 have been described at different positions, with the most prominent ones being R418W, R418G, R418Q and A726T (2,4,7). The A726T mutant, in which an alanine is replaced by a threonine residue (Figure 1) in the C1b domain, influences ADCY5’s flexibility. This single-point mutation is discussed to alter the structure of the ATP-binding pocket by affecting the interaction strength between C1 and C2 domains (2,3). The most abundant ADCY5 mutation is R418W (Figure 1) where a positively charged amino acid with a relatively long aliphatic chain (arginine) is replaced by a hydrophobic, aromatic amino acid (tryptophan). This single-point mutation is discussed to alter response as well as transmission of β-adrenergic receptor stimulation (2).

Gain-of-function ADCY5 mutants cause a significant increase in the catalytic hydrolysis of ATP as response to adrenergic stimulation (4,8), resulting in increased intercellular cAMP concentrations in striatal cells (8). All ADCY5 mutations result in kinetic disorders or dyskinesia that effect all muscles (2,4,9). A combination of linkage analysis and exon sequencing showed a strong evidence for mutations of ADCY5 being responsible for Familia Dyskinesia and Facial Myokymia (FDFM) (2,3).

The inhibition of ADCYs impacts the conversion of ATP to cAMP in the gain-of-function mutants and therefore reduces intracellular cAMP concentrations (10,11). Increasing evidence suggests that A_2A_ receptor antagonists exert positive effects on the symptoms of ADCY5-related dyskinesia (8). Caffeine is an A_2A_ antagonist and is hypothesized to alter cAMP levels in striatal neurons (12). Theophylline showed a higher potency regarding different metabolic effects compared to caffeine (13), as well as improved ergogenic effects during whole body exercise (13). Very recently, a report has been published, giving insights into the treatment of ADCY5-related dyskinesia patients with caffeine and describing the effects on frequency and duration of paroxysmal movement disorders, baseline movement disorders, and other motor and no-motor features (14). Overall, a consistent quality-of-life improvement was reported in 87% of patients. Only three out of 30 patients of this retrospective study reported a worsening of symptoms (14). Based on these findings, the use of caffeine has been suggested as first-line treatment of ADCY5-related dyskinesia (14).

In this work, we compare the effects of the three purines, caffeine, theophylline, and istradefylline, regarding their reduction of cAMP levels in ADCY5-overexpressing cell lines (ADCY5 wild-type and R418W mutant). We give a rational for administering theophylline to ADCY5-related dyskinesia patients based on the highly promising results obtained for one patient. We envision that theophylline has the potential to complement or even replace the therapy with caffeine for treating patients with ADCY5-related dyskinesia.

## Methods

### Materials and Methods

### Chemical and Reagents

All chemicals and reagents were obtained from Roth and Sigma Aldrich at the highest purity available.

### Cell Culture

HEK293T cells were stably transfected with vectors encoding GFP (for comparative purposes only), ADCY5 wild-type (ADCY5wt), and ADCY5 R418W mutant (ADCY5mut). Cells were cultured in Dulbecco’s modified Eagle’s Medium (DMEM) supplemented with 10% (v/v) fetal bovine serum (FBS) for two days in six-well plates at 37°C and 5% CO_2_; 4.4 × 10^5^ cells were cultivated in one well. Each treatment was performed in triplicate. Cells were incubated with caffeine, theophylline, and istradefylline at 37°C and 5% CO_2_ using different concentrations (caffeine and theophylline: 1 µM, 10 µM, 100 µM, 1 mM; istradefylline: 1 nM, 10 nM, 100 nM, 1 µM) for different times (10, 30, 60, 120, 240, and 480 min). Time-course experiments were performed in six replicates. Cells were harvested with PBS buffer, washed, and centrifuged at 1,500 x*g* for 5 min. 200 µl of trizol reagent (9.5 g guanidinium thiocyanate, 3.1 g ammonium thiocyanate, 3.5 ml of 3 M sodium acetate, 5 g glycerol, 48 ml Roti Aqua-Phenol in 100 ml total volume) were added to the cell pellets for cell lysis and inactivation of cAMP degrading enzymes. Cells were centrifuged with 30-kDa molecular weight cut-off filters (Amicon Millipore) at 14,000.0 x*g* for 10 minutes. Filtrates were stored at 4°C before they were analyzed by LC-MS/MS.

### Liquid Chromatography-Tandem Mass Spectrometry (LC-MS/MS)

LC separation of nucleotides (cAMP, AMP, ATP, cGMP, GMP, GDP, and GTP) was performed with a UPLC I-Class FTN system (Waters) equipped with an Atlantis Premier BEH C18 AX column (2.1 mm x 50 mm, 1.7µm, Waters). Separation was performed at a flow rate of 400 µl/min with the LC gradient as indicated in Figure 4C; solvent A: 0.2% (v/v) formic acid in water, solvent B: 0.2% (v/v) formic acid in acetonitrile. The UPLC system was directly coupled to a Xevo TQ-XS mass spectrometer (Waters) with electrospray ionization (ESI) source. Multiple-reaction monitoring (MRM) was performed using specific transitions for the nucleotides cAMP, AMP, ATP, cGMP, GMP, GDP, and GTP.

## Results

GFP, ADCY5wt, and ADCY5mut cells were compared regarding the ADCY5-catalyzed cAMP production according to the workflow presented in Figure 2. Cells were incubated with caffeine, theophylline, and istradefylline at varying concentrations and samples were collected at different time points. All three drugs are purine derivatives that exert their effects by binding to the A_2A_ receptor, resulting in ADCY5 inhibition and reduced cAMP levels (Figure 1). As ADCY5-related dyskinesia is caused by single-point, gain-of-function ADCY5 mutations (Figure 1), the reduction of cAMP levels will result in an improvement of movement disorders. This effect of caffeine on ADCY5-related dyskinesia patients has been impressively described in a recent report (14).

**Figure 2.**
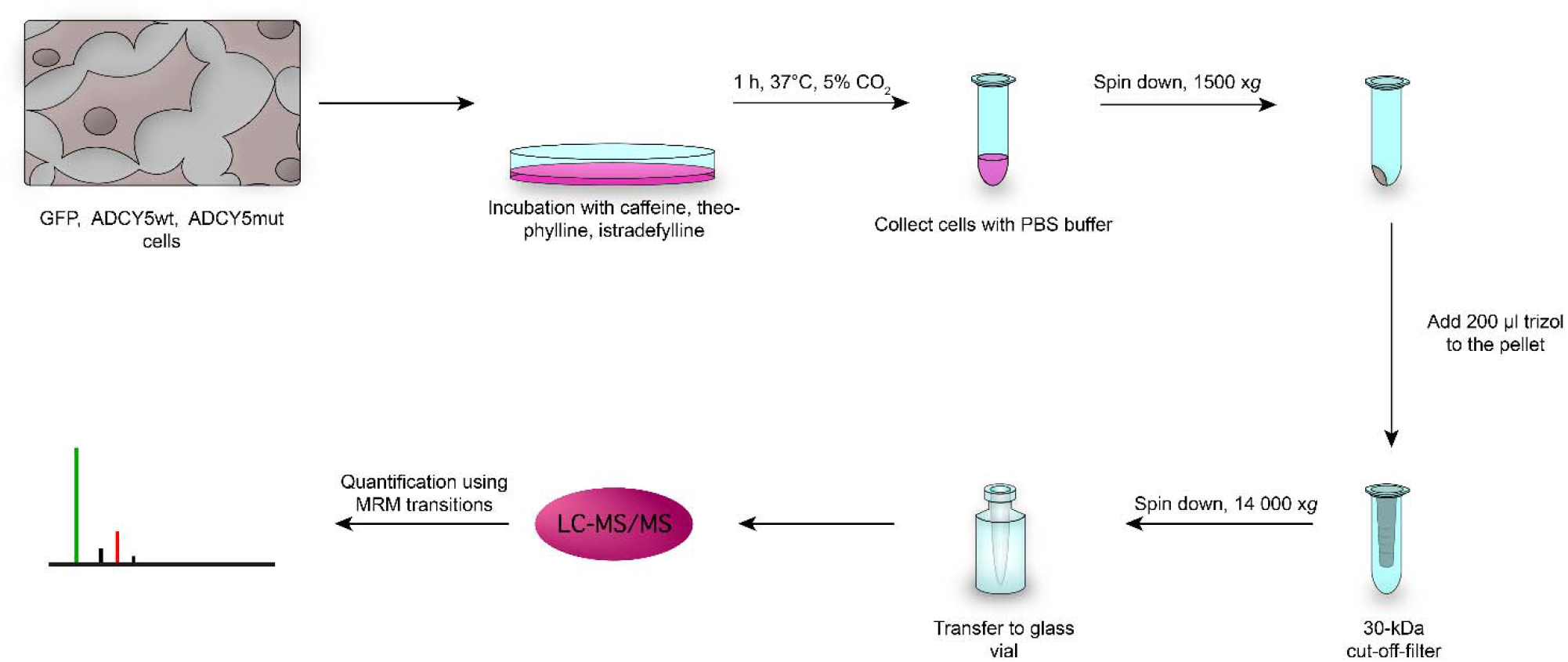
Analytical Workflow.

The main goal of the present study was to compare the effects of the three purines, caffeine, theophylline, and istradefylline, on cAMP concentrations at the cellular level. Caffeine, theophylline, and istradefylline differ in their K_i_ values (caffeine 23.4 µM, theophylline 1.7 µM, istradefylline 9.2 nM) at the A_2A_ receptor and should therefore exhibit different effects on the mediated ADCY5-catalyzed cAMP production (Figure 1). After harvesting and lysing the cells, filtrates were collected and analyzed by LC-MS/MS (Figure 2). Based on MRM analyses, cAMP concentrations were quantified after treatment with the three purine drugs using cells that stably express GFP (control), ADCY5 wild-type, and ADCY5 R418W mutant. This allowed us to gain detailed insights into the cAMP-reducing properties of caffeine, theophylline, and istradefylline at the cellular level.

### Comparison of caffeine, theophylline, and istradefylline regarding the reduction of cAMP levels

Consistent with the pivotal role of ADCY5 for cAMP production and its substantially increased catalytic activity caused by the R418W mutation, basal cAMP levels were upregulated ∼5-fold in ADCY5wt and ∼30-fold by ADCY5mut cells. Effects of the three purines were found to be most pronounced for ADCY5mut cells, showing a drastic reduction of cAMP levels already at concentrations of 100 µM caffeine and theophylline (Figure 3A). After ten minutes of drug treatment, initial effects on cAMP level reduction became visible (Figure 3B); after one hour of drug treatment the effects on lowering cAMP concentrations became even more pronounced (Figure 3C). Interestingly, cAMP concentrations that were observed after treatment of ADCY5mut cells with 100 µM theophylline were roughly at the same level as for non-treated ADCY5wt and GFP cells (Figure 3A). Apparently, treatment of ADCY5mut cells with a high concentration of theophylline results in basal cAMP levels that cannot be reduced any further. At the same time, rather large variations in cAMP levels were observed between different replicates in ADCY5mut cells (Figure 3A). This might be explained by the fact that the gain-of-function ADCY5 R418W mutant exhibits up to 30-fold increased basal cAMP levels compared to ADCY5 wild-type protein.

**Figure 3:**
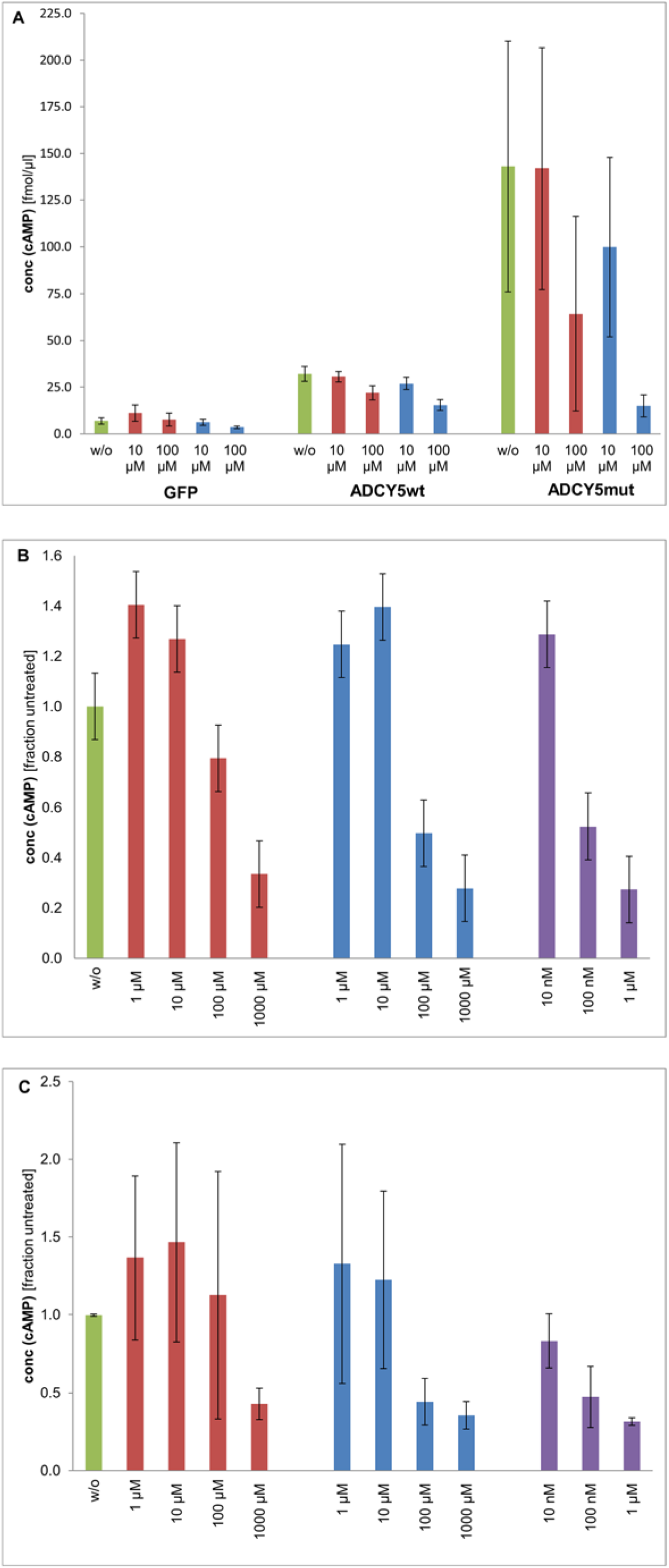
Influence of caffeine (red bars), theophylline (blue bars), and istradefylline (purple bars) at different concentrations on cAMP levels of different cell lines at different time points. Controls without drug treatment are shown as green bars. (**A**) GFP, ADCY5wt, and ADCY5mut cells, 1 h treatment. cAMP concentration of GFP cells (without ADCY5) are set to a value of 1.0. All cAMP concentrations of ADCY5mut cells are related to the GFP reference. (**B**) ADCY5mut cells, 10 min drug treatment (**C**) ADCY5mut cells, 1 h drug treatment. Experiments were performed in 3 replicates.

A comparison between the three purines under investigation revealed the most prominent reduction of cAMP concentrations in ADCY5mut cells for istradefylline, followed by theophylline and caffeine after already 10 minutes (Figure 3B). Apparently, the reduction of cAMP concentration correlates well with the respective K_i_ values (see above) of the three purines at the A_2A_ receptor. After one-hour treatment of ADCY5mut cells with caffeine, theophylline, and istradefylline (Figure 3C), the reduction of cAMP concentrations essentially exhibits the same profile as that observed at 10 minutes of drug treatment.

### Determination of cAMP concentration by LC-MS/MS

To determine ADCY5 activity, cAMP concentrations were determined by LC-MS/MS, quantifying different nucleotides by an LC-MRM-MS/MS approach to rule out any misassignments. Specific transitions were considered for cAMP, AMP, ATP, cGMP, GMP, GDP, and GTP (Figure 4A). For cAMP identification and quantification, the MS signal response showed a linear behavior in a concentration range up to 10 pmol/µl (Figure 4B). AMP, cAMP, GMP, cGMP reference compounds were used at a concentration of 100 fmol each, showing an LC baseline separation of all nucleotides with a total elution time of 3.5 minutes (Figure 4C). Based on our LC-MRM-MS/MS method, reliable and unambiguous cAMP quantification was performed using the cells’ supernatants (Figure 4D). As trizol reagent was used to stop all cellular reactions, phosphodiesterases that are eventually present will also be inhibited, resulting in a lack of cAMP-to-AMP conversion. We are therefore confident that the cAMP concentrations determined with our workflow (Figure 2) give a correct reflection of the enzymatic ADCY5 activity in the cellular systems studied herein.

**Figure 4:**
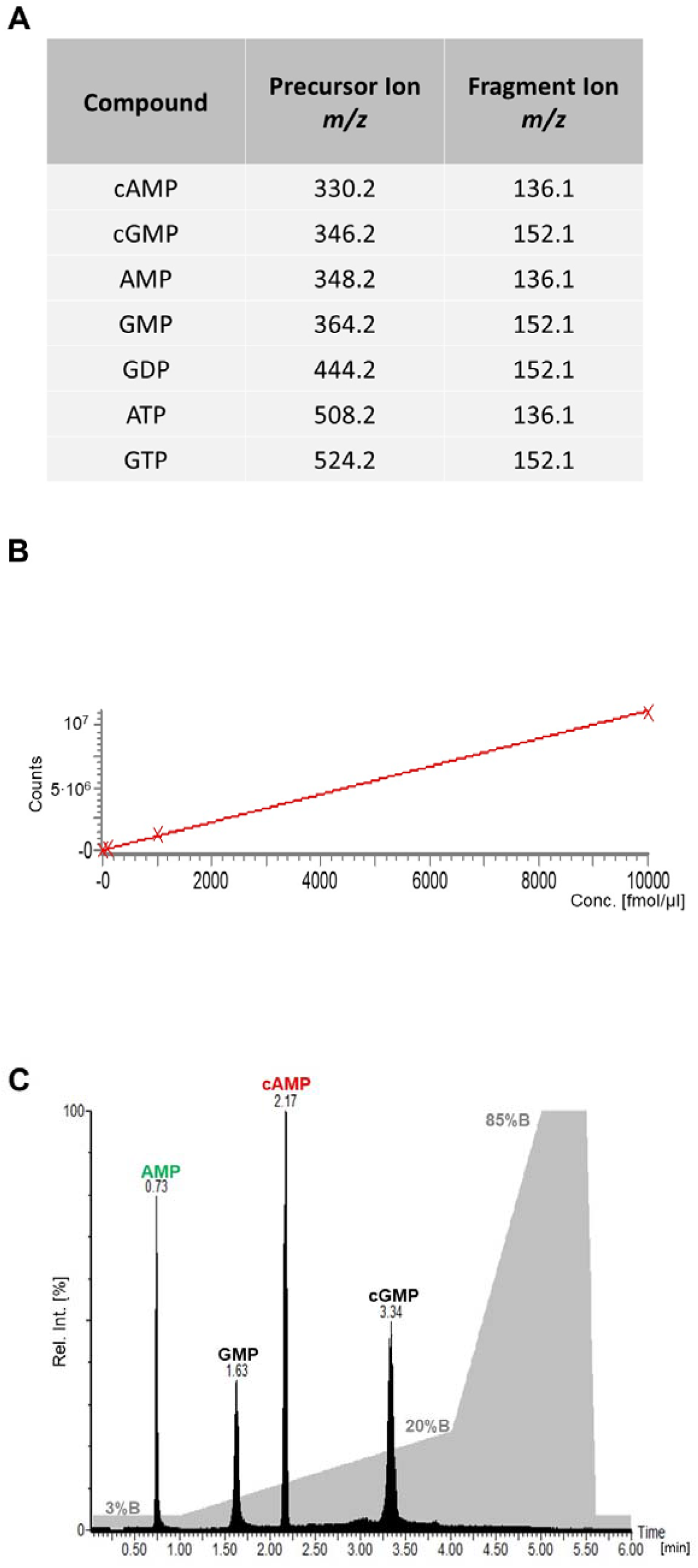
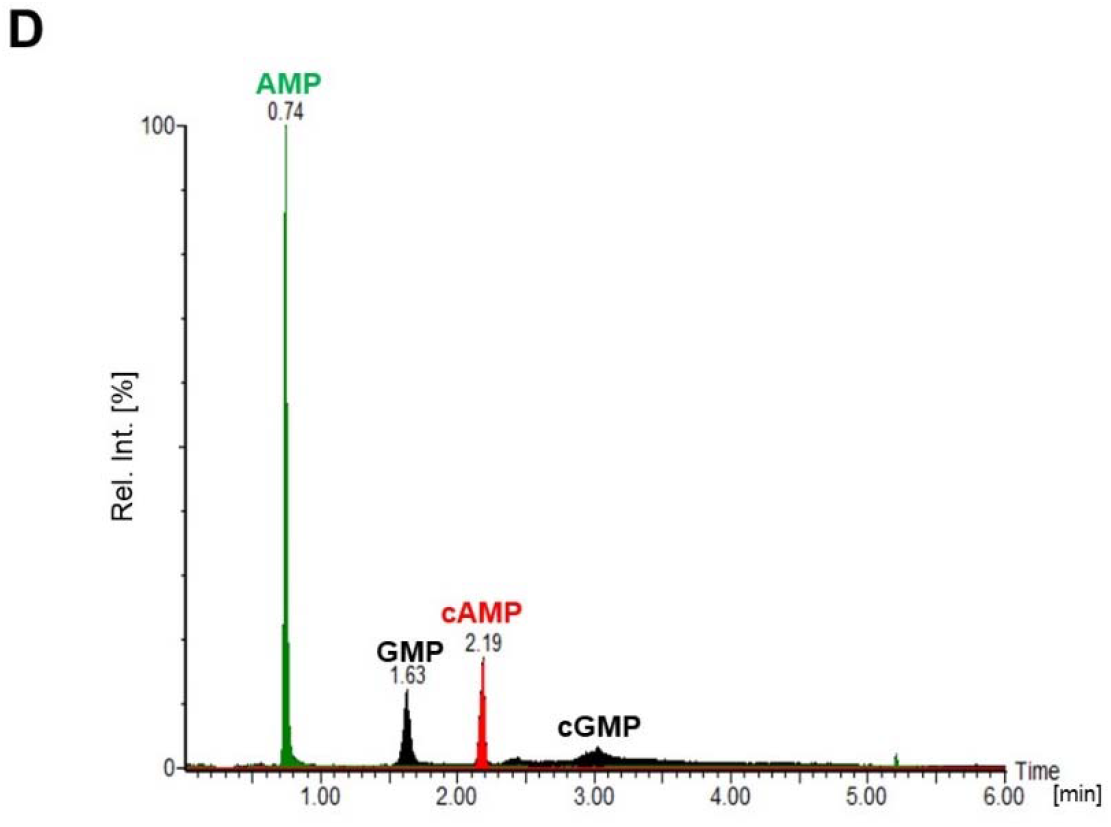
(**A**) Selected transitions in triple-quadrupole MS/MS measurements for selected nucleotides. (**B**) Linearity of mass spectrometric response for cAMP in the concentration range between 0-10 pmol/µl. (C) LC-MRM-MS/MS analysis (100 fmol) of four selected nucleotides (AMP, cAMP, GMP, cGMP). The TIC (Total Ion Current) of the LC elution is shown in black, the LC gradient is shown in grey (%B). (**D**) LC-MRM-MS/MS of supernatants (see Figure 2) for selected nucleotides.

### Time course of caffeine and theophylline treatment on ADCY5mut cells

Treatment of ADCY5mut cells with caffeine and theophylline (10 µM and 100 µM, each) over a time course of 480 minutes revealed a reduction of cAMP levels at both concentrations (Figure 5). Strikingly, the cAMP reduction was more pronounced for theophylline compared to caffeine.

**Figure 5:**
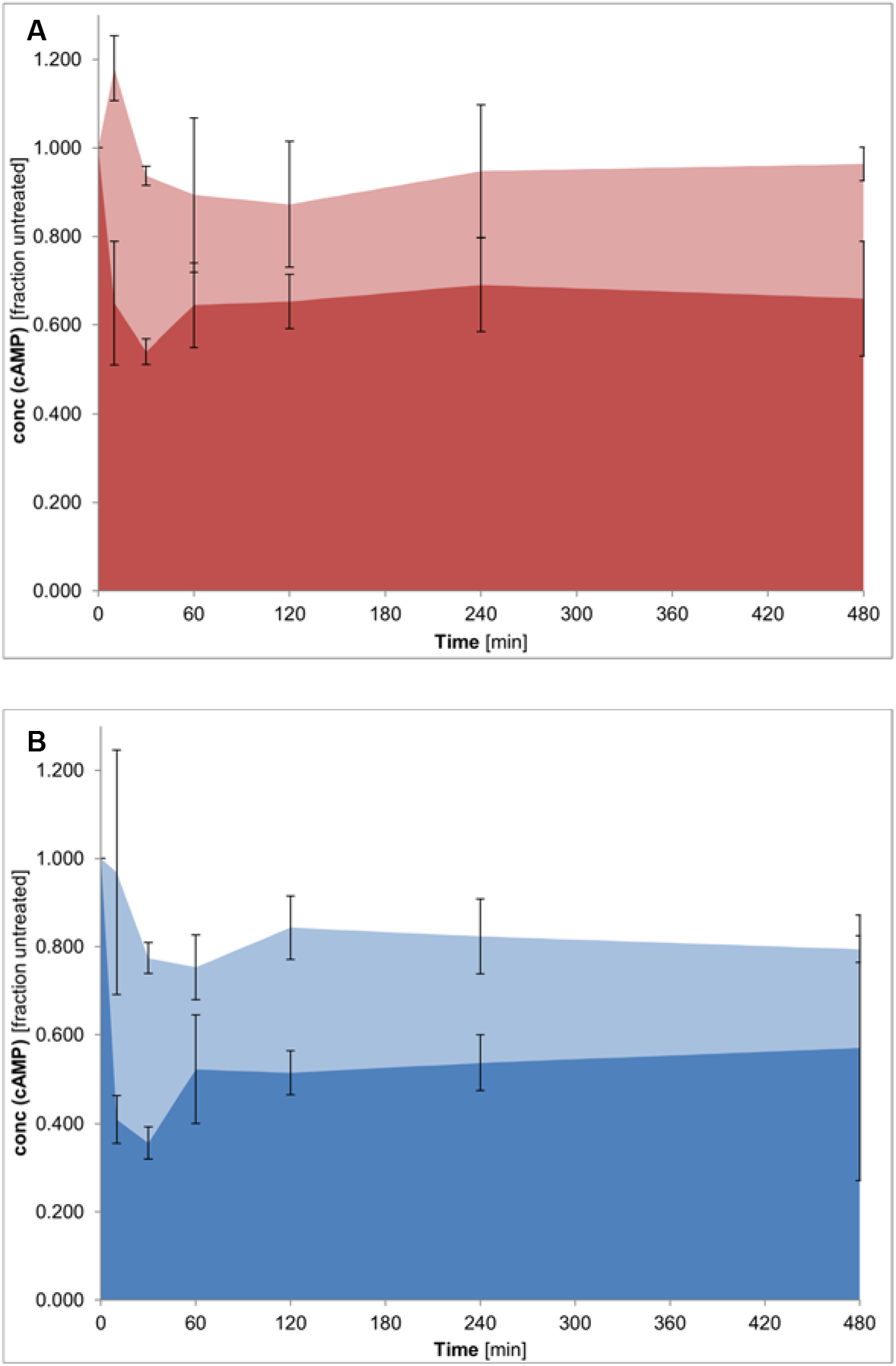
Time dependence of (**A**) caffeine (10 µM: light red, 100 µM: red) and (**B**) theophylline (10µM: light blue, 100µM: blue) treatment of ADCY5mut cell lines. cAMP concentrations were monitored over a time range of 8 hours. cAMP concentration of GFP cells (without ADCY5) are set to 1.0. All cAMP concentrations measured for ADCY5mut cells are related to the GFP reference. Experiments were performed in 6 replicates.

### Theophylline treatment of a patient with ADCY5-related dyskinesia

Given the promising results of reducing cAMP concentrations by theophylline, especially in ADCY5mut cells, we hypothesized that theophylline might be an alternative for caffeine to treat patients with ADCY5-related dyskinesia (14). Theophylline is available as slow-release formulation and has been shown to exhibit only minor side effects in children with status asthmaticus that were treated with low-dose theophylline (5-7 mg/kg per day) (15). Administering theophylline as slow-release formulation might be favorable compared to caffeine and result in a more efficient reduction of cAMP levels in patients (16) (17).

A preschool aged patient with ADCY5-related dyskinesia that had been successfully treated with 150 mg of caffeine (3x 50 mg daily) was treated with 400 mg of slow-release theophylline (2x 200 mg daily). Initially, theophylline and caffeine were given in combination, but in the course of the therapy, the daily caffeine dose was reduced over a period of five months until theophylline was administered alone. The theophylline dose was gradually increased from ∼6 mg/kg/day to ∼23 mg/kg/day. After 6-8 weeks, theophylline blood levels ranged between 7.5 mg/l to 21.1 mg/l.

Already at a theophylline dose of ∼12 mg/kg/day, divided into two single doses, the following effects were observed: The patient straightened up, showed increased muscle tonus, was able to stand independently, and walk six to ten free steps. The quality of sleep improved as dyskinetic movements subsided completely when falling asleep. At the same theophylline dose (∼12 mg/kg/day), the paroxysmal dyskinesia continued to fluctuate and recurrent infections of the patient led to a significant deterioration of every symptom, such as the loss of independent standing. Infection-associated deterioration of symptoms persisted despite increasing the daily dose of slow-release theophylline to ∼18 mg/kg/day.

By increasing the theophylline dose to ∼23 mg/kg/day, continuous improvements were observed. Supported by foot orthoses, the patient was able to walk ∼50 meters hand-held and 7 meters freely, and climbed stairs independently. The paroxysmal dyskinesia showed a decrease in frequency and duration, while theophylline had little effect on dysarthria and functional hypersalivation.

On a scale from 0 (no improvement) to 10 (no symptoms), theophylline had a more pronounced influence in the afternoon (6-7), while its influence was ranked between 3-4 in the morning. The duration and frequency of episodes cannot be reliably quantified as different causes (light bacterial or viral infections, stress, joy, etc.) influence motoric conditions during theophylline treatment.

We did not observe any adverse side effects during theophylline dose titration; arterial blood pressure, pulse rate, and theophylline-related laboratory parameters were within the given reference ranges. Even at the maximum dose of ∼23 mg/kg/day, sleep quality was greatly improved with no interruption of sleep for ten hours.

## Conclusions

We show that cAMP levels in ADCY5-overexpressing cells were reduced with all three purines, caffeine, theophylline, and istradefylline, under investigation. The effects were most prominent for the gain-of-function ADCY5mut cell line (R418W) and were in agreement with the Ki values of the three purines at the A_2A_ receptor: The most prominent reduction of cAMP levels was observed for istradefylline, followed by theophylline, while caffeine showed the lowest cAMP reduction. As istradefylline exhibits severe side effects even at low-dose application, it is not an approved drug in Germany (18). Theophylline, on the other hand, is available as slow-release formulation, showing only minor side effects even in pediatric applications. Theophylline was therefore administered as slow-release formulation to a preschool aged patient with ADCY5-related dyskinesia. Independent standing and walking as well as sleep quality, were substantially improved when treating the patient with theophylline compared to caffeine.

## Data Availability

All data produced in the present study are available upon reasonable request to the authors.

## Ethics Statement

The ethics committee of the University of Leipzig was informed and approved this study. The study has been registered at the DRKS (ID: DRKS00029154).

## Acknowledgements

AS acknowledges financial support by the DFG (RTG 2467, project number 391498659 “Intrinsically Disordered Proteins – Molecular Principles, Cellular Functions, and Diseases”, RTG 2751 “InCuPanC”, project number 449501615, INST 271/404-1 FUGG, INST 271/405-1 FUGG, and CRC 1423, project number 421152132), the Federal Ministry for Economic Affairs and Energy (BMWi, ZIM project KK5096401SK0), the region of Saxony-Anhalt, and the Martin Luther University Halle-Wittenberg (Center for Structural Mass Spectrometry). Dr. Wendy H. Raskind, University of Washington, Seattle, WA, is acknowledged for providing the ADCY5 plasmid. Dr. Frank Erdmann, Martin Luther University Halle-Wittenberg, is acknowledged for initial discussions.

## Author Contributions

DT, MK, and ML conducted experiments and analyzed the data; WFG and CA created the figures; AS and WFG wrote the manuscript; SH, AM, and AS supervised the study.

